# Comparison of the Sensitivity and Specificity of Commercial Anti-Dengue Virus IgG Tests to Identify Persons Eligible for Dengue Vaccination

**DOI:** 10.1101/2024.04.19.24306097

**Authors:** Freddy A. Medina, Frances Vila, Laura E. Adams, Jaime Cardona, Jessica Carrion, Elaine Lamirande, Luz N. Acosta, Carlos M. De León-Rodríguez, Manuela Beltran, Demian Grau, Vanessa Rivera-Amill, Angel Balmaseda, Eva Harris, Zachary J. Madewell, Stephen H. Waterman, Gabriela Paz-Bailey, Stephen Whitehead, Jorge L. Muñoz-Jordán

## Abstract

The Advisory Committee on Immunization Practices (ACIP) recommended that dengue pre-vaccination screening tests for Dengvaxia administration have at least 98% specificity and 75% sensitivity. This study evaluates the performance of commercial anti-DENV IgG tests to identify tests that could be used for pre-vaccination screening. First, for 7 tests, we evaluated sensitivity and specificity in early convalescent dengue virus (DENV) infection, using 44 samples collected 7-30 days after symptom onset and confirmed by RT-PCR. Next, for the 5 best performing tests and two additional tests (with and without an external test reader) that became available later, we evaluated performance to detect past dengue infection among a panel of 44 specimens collected in 2018-2019 from healthy 9-16-year-old children from Puerto Rico. Finally, a full-scale evaluation was done with the 4 best performing tests using 400 specimens from the same population. We used virus focus reduction neutralization test and an in-house DENV IgG ELISA as reference standards.

Of seven tests, five showed ≥75% sensitivity detecting anti-DENV IgG in early convalescent specimens with low cross-reactivity to Zika virus. For the detection of previous DENV infections the tests with the highest performance were the Euroimmun NS1 IgG ELISA (sensitivity 84.5%, specificity 97.1%) and CTK Dengue IgG rapid test R0065C with the test reader (sensitivity 76.2% specificity 98.1%). There are IgG tests available that can be used to accurately classify individuals with previous DENV infection as eligible for dengue vaccination to support safe vaccine implementation.

## Introduction

The mosquito-borne dengue virus (DENV) is a growing threat globally. Approximately half of the world’s population lives in at-risk areas, and there are almost 400 million infections annually (1, 2). There are four related, but antigenically distinct DENV serotypes that can produce symptomatic disease in approximately 25% of infections. Individuals with dengue can experience a range of symptoms, from mild manifestations to severe disease that requires hospitalization and can result in death. Infection with one DENV serotype is thought to provide lifelong immunity against the infecting serotype. After a primary DENV infection, the antibody levels wane after 6-9 months to a level where they remain stable for years, and the risk of developing severe illness during a second infection with any of the other three serotypes increases (3). The risk of severe dengue decreases for the third and fourth DENV infections.

Implementing Dengvaxia^®^, the first licensed dengue vaccine is complicated by the necessity of confirming a previous dengue infection prior to vaccination. In 2019, the Food and Drug Administration (FDA) approved Dengvaxia^®^ for individuals aged 9-16 years who have a laboratory-confirmed previous DENV infection and live in endemic areas of the United States. After careful review, the Centers for Disease Control and Prevention (CDC) Advisory Committee on Immunization Practices (ACIP) recommended the use of Dengvaxia^®^ in 2021, and provided guidance for its implementation in the United States and its territories and freely associated states (4). Tests to be used for determining previous dengue infections prior to vaccination must have a minimum of ≥ 75% sensitivity and ≥ 98% specificity, with a neutralizing antibody titer at 50% inhibition (NT50) as the reference standard for the United States (4, 5). These criteria were based on evidence that although a previous DENV infection can be demonstrated by laboratory-confirmed RT-PCR detection or a positive NS1 test during the acute phase of infection, a large proportion of DENV infections are asymptomatic or mildly symptomatic and may not have been tested or records of previous testing may not have been maintained. Evidence of a previous DENV infection by serological testing for anti-DENV IgG antibodies is the most feasible recommendation but can be problematic due to the cross-reactivity between DENV and other flaviviruses such as Zika virus (ZIKV), which circulated in many dengue-endemic areas. Although there are many commercial tests for the detection of anti-DENV IgG, none of them are currently licensed by the FDA, and most were developed for diagnostic purposes during the early convalescent phase of infection, when antibody levels are high, rather than for determining serostatus years after infection when antibody levels are lower. Moreover, many of the currently available commercial anti-DENV IgG tests were developed prior to the emergence of ZIKV and little is known about the level of flavivirus cross-reactivity that occurs in these tests. A systematic review in 2019 did not identify studies describing the evaluation of lateral flow tests for the detection of anti-DENV IgG long after (≥ 1 year) infection that could indicate a potential use of these tests for pre-vaccination screening (6). Since then, limited publications describe evaluations of tests for the purpose of pre-vaccination screening; but studies using side-by-side comparisons of commercial tests with specimens from typical eligible healthy candidates are lacking (7-11). Comparative studies are needed to determine the best commercial anti-DENV IgG tests to guide dengue vaccine implementation efforts in the United States.

Our aim in this study was to determine the sensitivity and specificity of commercial anti-DENV IgG tests that could be used to accurately classify individuals with previous DENV infection (PDI) as eligible for dengue vaccination in the United States as recommended by the ACIP. Serum specimens used for comparisons were obtained from Puerto Rico, the jurisdiction with the largest population at risk for dengue transmission in the United States. The specimens were from two sources: early convalescent, well-characterized specimens from people with DENV or ZIKV confirmed by RT-PCR or negative for both viruses, and samples from healthy 9–16-year-old children whose immune status was characterized by the combined results of a DENV/ZIKV focus reduction neutralization test (FRNT) and the CDC DENV IgG ELISA.

## Methods

### Participant enrollment and ethics statement

Early convalescent specimens were retrospectively obtained from the Sentinel Enhanced Dengue and Acute Febrile Illness Surveillance System (SEDSS), as described previously (12, 13). Samples to evaluate test performance for previous dengue infections were retrospectively obtained from the Communities Organized to Prevent Arboviruses (COPA) project in Puerto Rico that conducts annual serosurveys among participants 1-50 years of age. Participants were recruited into COPA as previously described with written informed consent or assent obtained from all study participants according to a protocol approved by Institutional Review Boards at the Centers for Disease Control and Prevention (CDC) and Ponce Medical School Foundation, Inc (PMSF). We used random samples among non-ill children 9-16 years of age collected in 2018 and 2019 (14). Specimens from primary ZIKV infections were obtained from a pediatric cohort in Nicaragua collected in 2018 and 2019 (15). The ZIKV infections were confirmed by real-time RT-PCR during the acute episodes in 2016, and absence of DENV infections was confirmed in paired annual samples tested with a DENV inhibition ELISA (16).

### Test selection

A search was performed for commercial tests that detect anti-DENV IgG. A total of 60 tests (46 ELISA and 14 rapid diagnostic tests [RDT]) from 34 manufacturers were identified. This group was narrowed down to 23 tests (11 ELISAs, 12 RDTs) by selecting only tests that reported performance data on their website or in their instructions for use. All but two tests reported greater than 90% specificity. The tests with sensitivity lower than 75% were removed from consideration. This group was further reduced to 7 tests (5 ELISAs, 2 RDTs) since some manufacturers or representatives were not reached after multiple attempts to contact them by both e-mail and telephone (n=9), or if the manufacturer stated that the product was withdrawn from sale (n=5). The 7 tests selected were included in the first phase of the evaluation to determine performance in early convalescent specimens, as these tests have been manufactured for use in disease diagnosis. Based on their performance, 5 of these tests were selected for further analysis. During this evaluation, two versions of a rapid test manufactured by CTK Biotech became available for the detection of IgG antibodies as an indication of prior DENV infection in asymptomatic individuals. CTK Biotech also offers the Alta rapid test reader (RTR-1) as an optional tool to automate and standardize the presence or absence of test line bands. Therefore, 7 tests moved to another round of the evaluation using a panel of samples from patients with confirmed previous dengue infection: the 5 tests with highest accuracy selected from the evaluation with early convalescent specimens and the 2 CTK test modalities. The 4 best performing tests were further evaluated for detection of previous dengue infection in a survey of healthy individuals. The tests used in these evaluations were purchased directly from the manufacturers, stored at the recommended temperatures, and used prior to their expiration dates. The 9 tests evaluated are listed in Table 1.

**Table 1:**
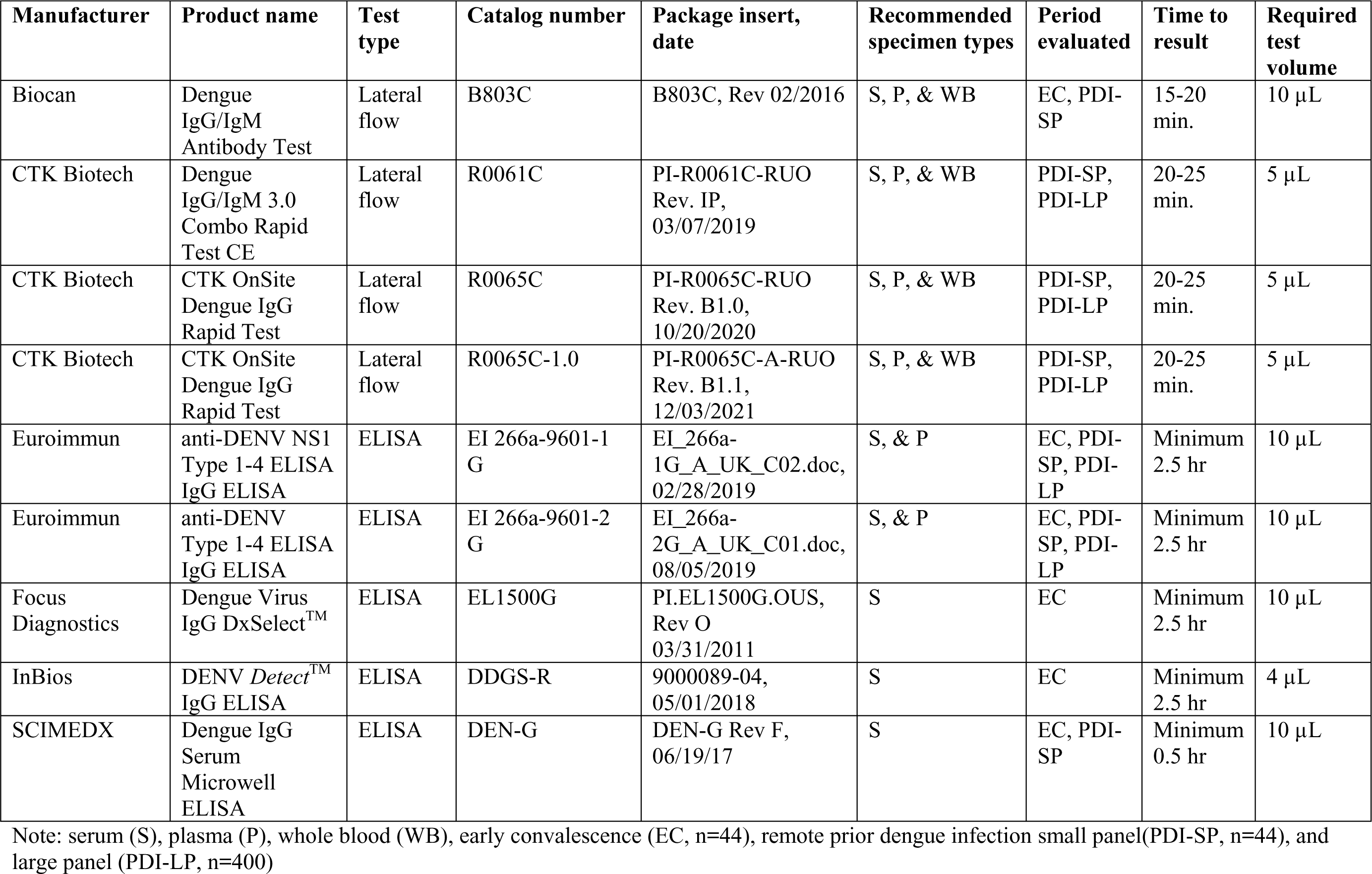
Commercial anti-DENV IgG diagnostic tests evaluated.

### Focus reduction neutralization test (FRNT)

FRNT assays for DENV-1-4 were performed blinded at the Laboratory of Viral Diseases, National Institute for Allergy, and Infectious Disease (NIAID), National Institutes of Health (NIH) as previously described (17). Viruses used in the assay included: DENV-1 Western Pacific (Nauru/74), DENV-2 NGC Prototype (1944), DENV-3 Sleman/78, and DENV-4 814669 (Dominica/81). Serum specimens were used at dilutions of 1:5, 1:20, and 1:80 with antibody titers defined as the highest serum dilution that resulted in >50% reduction (FRNT50) in the number of immunostained virus foci. A specimen was considered positive for neutralization against the infecting serotype when the FRNT titer was >4 for DENV-1, >15 for DENV-2, >15 for DENV-3, and >4 for DENV-4 (Supplemental Figure S1). Specimens with no neutralizing antibodies against a virus were given a value of 4. We used the FRNT50, considered the gold standard for measuring neutralizing antibodies against dengue virus and other flaviviruses, in combination with the CDC DENV IgG test as our reference standard.

### CDC DENV IgG ELISA

The CDC DENV IgG assay was calibrated based on the NIH FRNT50 assay and a modified version of Miagostovich *et. al* (18). Capture antibody (4G2 Mouse anti-Flavivirus Envelope Protein, Native Antigen Company, Oxford, United Kingdom) was diluted 1:6,000 in 0.1 M carbonate–bicarbonate buffer pH 9.6 and plates were incubated overnight at 4°C. Plates were washed three times in phosphate buffered saline (PBS) and incubated with blocking buffer (PBS pH 7.4/0.05% Tween 20/3% normal goat serum) for 1 hour at 37°C. After removing blocking buffer, a DENV-1-4 antigen mix (DENV-1-4 virus-like particle [VLP] recombinant antigens) titrated between 0.1 ng/µl-0.7 ng/µl for each antigen, Native Antigen Company, Oxford, United Kingdom) diluted in blocking buffer was added to each well and incubated covered for 1 hour at 37°C. Plates were washed three times in PBS. Each plate contained a high positive control, low positive control, high negative control, two low negative controls, and a cutoff control. Serum specimens, positive controls, negative controls, and a cutoff control were diluted 1:100 in a solution of PBS pH 7.4 containing 3% non-fat dry milk/ 0.05% Tween-20 and tested in duplicate. The positive and cutoff controls were created using pooled early convalescent specimens collected 7–30 days post-onset (DPO) of symptoms from RT-PCR confirmed DENV-1–4 cases and diluted in PBS. Plates were washed three times in PBS and a horse radish peroxidase conjugated anti-human IgG diluted in a solution of PBS pH 7.4 containing 3% non-fat dry milk/ 0.05% Tween-20 was added and incubated in the dark for 1 hour uncovered at room temperature. Plates were washed six times with PBS and ABTS substrate added to each well. After a 60-minute room temperature incubation in the dark, plates were read in a microplate spectrophotometer at 405 nm without a blank. Optical density (OD) values and antibody index ratio (AIR) calculations for the controls and samples are needed for test run validation and interpretation of results, accordingly.

AIR = ([control or sample OD / cutoff control OD] *10) Positive: AIR ≥ 15

Equivocal: AIR 10-14.99

Negative: AIR <10

Non-concordant results were repeated twice by CDC DENV IgG ELISA and FRNT50 to confirm results. All specimens except one were concordant in both assays.

### ZIKV EDIII IgG ELISA

The ZIKV EDIII IgG ELISA was performed as previously described in Adams et. al. with the modification of using diluted human monoclonal antibody ZKA190 as a cutoff control for inter-plate consistency (19). High-binding microtiter 96-well plates (Greiner Bio-One) were coated with streptavidin diluted to 4 μg/mL in Tris-buffered saline (TBS, pH 7.4), and incubated for 1 hour at 37°C to ensure optimal binding. After incubation, the plate was decanted, washed thrice with wash buffer (TBS containing 0.2% Tween 20). To minimize non-specific binding, the plate was blocked using a blocking solution (3% milk in TBS containing 0.05% Tween 20) for 1 hour at 37°C. Biotinylated EDIII antigen diluted at a concentration of 2 μg/mL in blocking buffer was captured in plates after a 1-hour incubation at 37°C. Plates were washed thrice with wash buffer. After removal of the blocking solution, serum samples at a 1:20 dilution in blocking buffer were added to each well and incubated for 1 hour at 37°C for binding of ZIKV-specific IgG antibodies to the captured EDIII antigen. Following another series of three washes, goat anti-human IgG alkaline phosphatase-conjugated secondary antibody (Sigma) was added at a 1:2,500 dilution and incubated for 1 hour at 37°C. After a final washing, SigmaFast AP substrate was added, and the reaction was developed in the dark for 30 minutes at room temperature. The plate was then read at a 405 nm wavelength setting without a blank. For validation and interpretation of results, optical density (OD) values and antibody index ratio (AIR) calculations were necessary for controls and samples.

AIR = ([control or sample OD / cutoff control OD] *10)

Positive: AIR > 10

Negative: AIR ≤10

## Laboratory testing

### Early convalescent specimens

A serum panel was assembled with early convalescent specimens (EC panel, N=44) collected 7– 30 (average = 12) days post-onset (DPO) of symptoms from cases in a convenience series within SEDSS. These cases were confirmed as either DENV or ZIKV positive or negative by RT-PCR on the first specimen collected from each case (DPO 0-5) and whose immune status was determined based on testing a convalescent sample with the CDC DENV IgG or ZIKV IgG assays, as previously described (19). Unexposed specimens (n=8) were negative in the CDC Trioplex RT-PCR, negative for DENV and ZIKV IgG, and IgM negative in the acute and early convalescent specimens for DENV and ZIKV. Specimens from ZIKV RT-PCR positive cases with negative and DENV IgG negative results were named primary ZIKV (n=14). DENV primary specimens (n=12) were DENV RT-PCR-positive and DENV and ZIKV IgG-negative. DENV secondary specimens (n=8) were DENV RT-PCR positive, negative for ZIKV IgG, and positive for DENV IgG.

### Previous DENV infection specimens

The following immune status classification criteria were established for samples from a serosurvey of asymptomatic 9-16 Y/O children (COPA) that were available for this study: Specimens were classified as positive for DENV only if they were positive in the DENV FRNT and CDC DENV IgG ELISA. Specimens that neutralized a single DENV serotype and were not positive in the ZIKV EDIII IgG ELISA were classified as monotypic DENV, and specimens that neutralized two or more DENV serotypes and were not positive by ZIKV EDIII IgG ELISA were classified as multitypic DENV. Specimens that did not neutralize any DENV serotype and were ZIKV EDIII IgG ELISA positive were classified as primary ZIKV. Specimens that neutralized any DENV serotype and were positive in the ZIKV EDIII IgG ELISA were classified as multi-flavivirus. Specimens that did not neutralize DENV nor tested ZIKV EDIII IgG ELISA positive, and specimens that were only positive in the CDC DENV IgG ELISA or FRNT50 were classified as unexposed.

We constructed a panel with 44 specimens selected out of from the total COPA serosurvey to assess previous DENV infections (PDI). The 44 samples were classified according to the above-stated immune status classification criteria for asymptomatic cases with the following proportions: unexposed (n=8), primary ZIKV (n=14), monotypic DENV (n=13) and multitypic DENV (n=9). Multi-flavivirus samples were not included in this panel. We then expanded our evaluation to a panel of 400 samples from the COPA serosurvey, which included a blinded selection of 50 specimens from each year of age (9–16), within a total of 750 participants of that age group. These specimens were classified according to the same immune status clarification criteria (Table 2). FRNT50 titers for the 400 samples for each DENV serotype according to their immune status classification can be found in supplementary Figure S1.

**Table 2:** Immune status classification of 400 specimens used to evaluate use of DENV IgG tests to determine PDI.

| **Immune Status** | **NIH FRNT50**<br>**DENV** | **ZIKV EDIII**<br>**IgG ELISA** | **CDC DENV**<br>**IgG** | **TOTAL = 400** |
| --- | --- | --- | --- | --- |
| Monotypic DENV | POSITIVE<br>(one serotype) | NEGATIVE | POSITIVE | DENV-1= 16<br>DENV-2= 12<br>DENV-3= 4<br><u>DENV-4= 4</u><br>ALL= 36 |
| Multitypic DENV | POSITIVE<br>($\geq 2$ serotypes) | NEGATIVE | POSITIVE | 77 |
| Primary ZIKV | NEGATIVE | POSITIVE | ANY<br>RESULT | 56 |
| Multi-flavivirus | POSITIVE<br>($\geq 1$ serotype) | POSITIVE | POSITIVE | 80 |
| Unexposed | NEGATIVE | NEGATIVE | NEGATIVE | 150 |
| Unexposed | POSITIVE | NEGATIVE | NEGATIVE | 0 |
| Unexposed | NEGATIVE | NEGATIVE | POSITIVE | 1 |

### Additional evaluation for cross-reactivity with ZIKV specimens

To ensure that the tests with high performance for pre-vaccination screening with Puerto Rico specimens had little cross-reactivity with ZIKV, we tested an additional 22 well-characterized RT-PCR-confirmed past ZIKV infections from a cohort in Nicaragua and an additional 41 specimens from unexposed healthy individuals from Puerto Rico, both obtained from cases of a convenience series. In addition, 12 challenging samples from primary ZIKV positive patients were selected for their high ZIKV neutralizing and binding antibody levels. The specimens were collected at 3-4 months after RT-PCR confirmed infections in 2016-2017.

All serum specimens used in these evaluations were stored frozen after arrival to the CDC Dengue Branch, thawed and prepared in aliquots for virus neutralization, CDC DENV IgG ELISA, or commercial assay testing.

### Specimen testing

Specimen testing was conducted contemporaneously with the index tests and reference standards to ensure consistency and reliability in the results. All tests were performed and interpreted according to the manufacturer’s instructions for use and all specimens were tested in index tests and the reference standard. There was no missing data. ELISA equivocal/borderline/weakly reactive results were considered negative for calculations of test performance. Testing was performed by the same laboratory technician for all ELISA assays. Rapid tests were independently interpreted visually by two laboratory technicians who were blinded to specimen classifications. The CTK OnSite Dengue IgG Rapid Tests were also read with the CTK Alta Rapid Test Reader RTR-1 optimized for visual readout with the large panel (PDI, n=400) collected from healthy children 9-16 years of age in Puerto Rico. There were no adverse events associated with performing the index tests or the reference standard. This study was carried out and documented following the Standards for Reporting Diagnostic Accuracy Studies (STARD) guidelines (20). Participant flow and outcomes for the index tests are depicted using STARD flow diagrams (Figures S2-12). This study was conducted and reported in accordance with the Standards for Reporting Diagnostic Accuracy Studies (STARD) guidelines. The flow of participants for the index tests and their results was presented using STARD diagrams.

## Sample Size Calculation

The sample size for our diagnostic accuracy of previous DENV infection specimens was calculated based on the expected prevalence (45%) of Dengue in Puerto Rico and for the diagnostic tests expected to achieve a sensitivity of at least 75% and a specificity of 98% with a margin of error of ±3%. The significance level (α) was set at 0.05. Using the previously described parameters for calculations, the minimum sample size required was 374.

## Data analysis

### Sensitivity

Test sensitivity was measured as the evaluated test’s ability to correctly identify specimens that are positive for DENV IgG antibodies through two refence standards: the CDC DENV IgG ELISA and the DENV FRNT50 assays. A specimen was confirmed as DENV IgG positive if it tested positive in both assays serving as reference standards. Sensitivity was calculated as the number of anti-DENV positive specimens correctly detected by the test under evaluation, divided by the total number of anti-DENV positive specimens confirmed by CDC DENV IgG ELISA and classified as DENV immune by FRNT50, multiplied by 100.

### Specificity

Test specificity was measured as the evaluated test’s ability to correctly identify specimens that were not exposed to DENV (negative for DENV IgG antibodies) or were exposed to ZIKV only (positive for ZIKV IgG antibodies but negative for DENV IgG antibodies). For this evaluation, we considered non-concordant specimens—those that test positive in either the CDC DENV IgG ELISA or the DENV FRNT50 assay but not in both—as negative for DENV IgG antibodies. Specificity was calculated by adding the number of specimens that the evaluated tests correctly identified as negative (including true negative, equivocal/borderline/weakly reactive and non-concordant specimens) and only ZIKV exposed, divided by the total number of specimens classified as ZIKV primary (based on a positive result in the ZIKV EDIII IgG ELISA and a negative result in the DENV FRNT50) or DENV unexposed (based on negative results in both the CDC DENV IgG ELISA and the FRNT50), multiplied by 100.

### Confidence interval

The 95% confidence interval (CI) of the estimated sensitivity and specificity was calculated using the formula: P±1.96√P(1−P)/N, where P is the sensitivity or specificity, and N is the number of specimens tested. If the sensitivity or specificity was 100% it was substituted with 99.9% for P in the equation.

## Results

### Performance on early convalescent specimens

Seven tests were first evaluated for their capacity to detect anti-DENV IgG and their potential to minimize detection of cross-reactive ZIKV IgG in early convalescent specimens (n=44) (Table 1). All (7/7) tests had at least overall moderate (75%) to high (100%) sensitivity for anti-DENV IgG (Figure 1). The detection of anti-DENV IgG from secondary DENV was universal for all tests; but differences in test sensitivity were observed for primary DENV. Sensitivity for anti-DENV IgG in primary DENV specimens was lower in RDTs in comparison with ELISA tests. The RDTs detected over half of the primary DENV specimens. ELISA tests displayed high sensitivity for anti-DENV IgG in primary specimens, except for the SCIMEDX Dengue IgG serum microwell ELISA, which had a sensitivity comparable to RDTs. Despite the high sensitivity observed in the Focus DENV IgG DxSelect and InBios DENV Detect IgG ELISAs, they were excluded from further evaluation due to the high (50-57 %) reactivity observed in primary ZIKV specimens. Cross-reactivity in the remaining tests was low (7-14%) and warranted further evaluation. No test had false positives in specimens from unexposed individuals.

**Figure 1.**
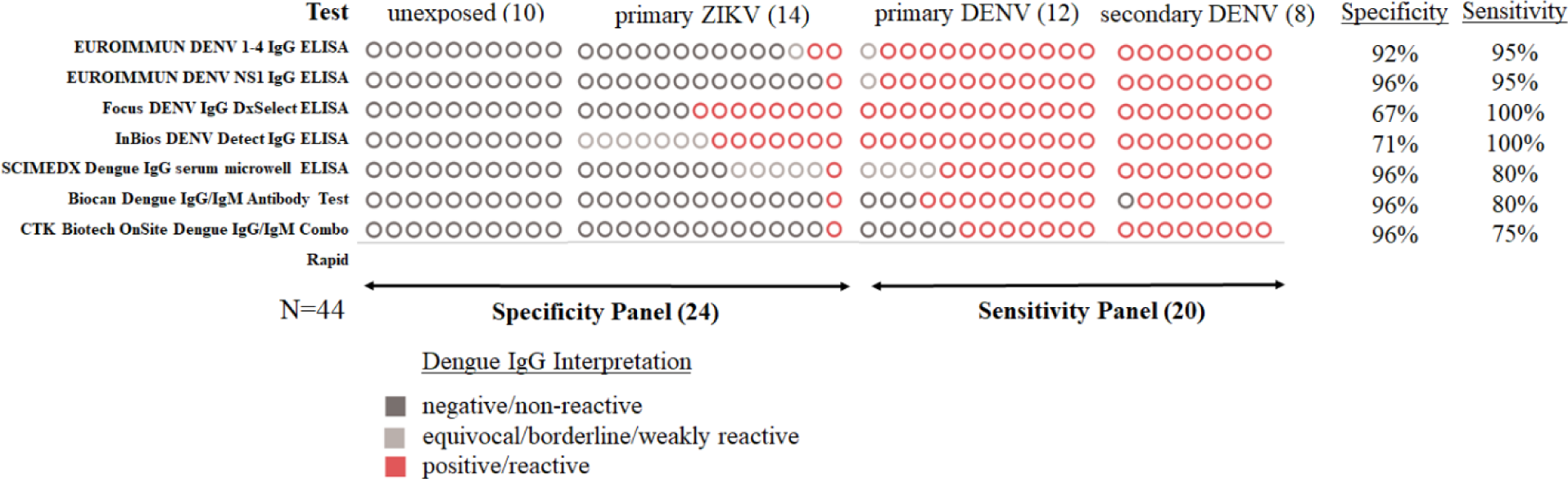
Comparison of anti-DENV IgG commercial tests evaluated for specificity and sensitivity with specimens collected during early convalescence. Note: Circles represent the number of specimens tested and are not in the same order by column.

### Performance to detect previous dengue infections with a small panel of 44 specimens

The 44-sample panel of specimens from non-ill children was used to assess 7 commercial anti-DENV IgG tests for their performance to detect previous dengue infections (PDI-SP, n=44); which included 5 tests that passed the EC panel and 2 IgG tests from CTK Biotech that became commercially available during the course of our evaluation (Table 1). The tests with the highest performance (specificity and sensitivity, respectively) were the Euroimmun DENV-1-4 IgG ELISA (86%, 86%), the Euroimmun DENV NS1 IgG ELISA (95%, 68%), the CTK Onsite Dengue IgG Rapid Test R0065C (95%, 82%) and the CTK Onsite Dengue IgG Rapid Test R0065C 1.0 (95%, 68%) (Figure 2). The sensitivity of Biocan Dengue IgG/IgM Antibody Test (27%), CTK Biotech OnSite Dengue IgG/IgM Combo Rapid (0%), and SCIMEDX Dengue IgG serum microwell ELISA (23%) was overall very low and not suitable for pre-vaccination screening. The test with the highest ZIKV cross-reactivity was the Euroimmun anti-DENV Type 1-4 ELISA IgG ELISA, which uses VLP antigen and had 3/14 primary ZIKV specimens test positive.

**Figure 2.**
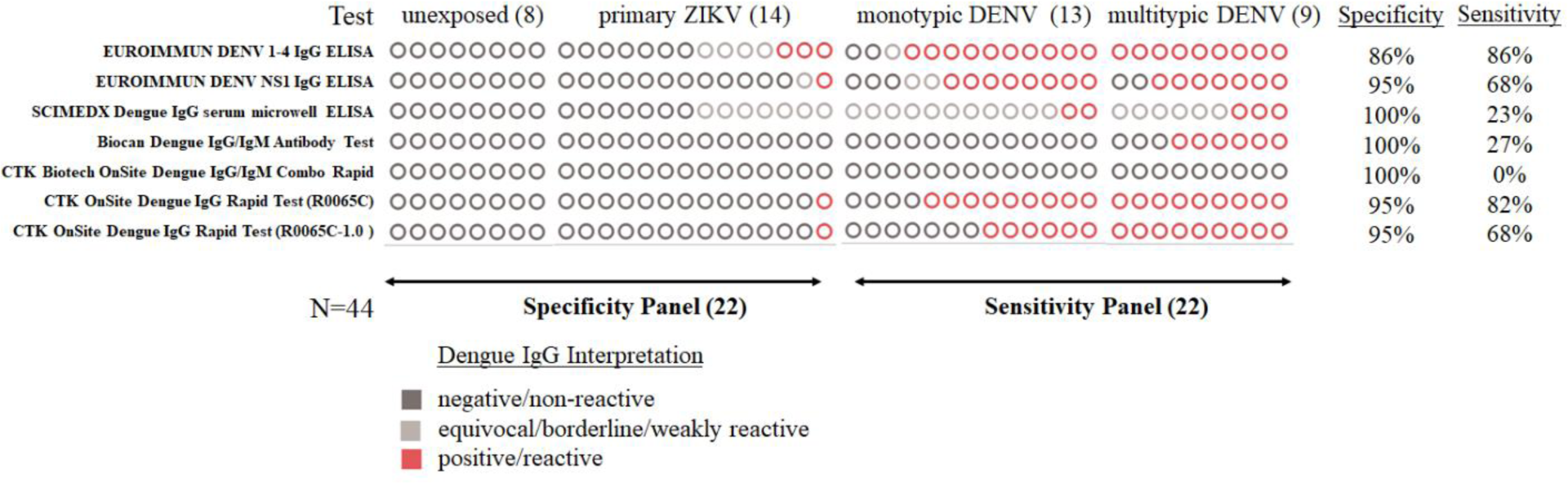
Comparison of anti-DENV IgG commercial tests evaluated for specificity and sensitivity of previous dengue infection in serum specimens collected from non-ill children (9-16 years of age). Note: Circles represent the number of specimens tested and are not in the same order by column.

### Performance to detect previous dengue virus infection with a community survey of 400 specimens

The full-scale evaluation was done with the two Euroimmun ELISAs and the two CTK OnSite Dengue IgG rapid tests. From the 750 children (age 9-16) in the COPA cohort, 400 specimens were selected for the test evaluation and categorized by immune status using the DENV/ZIKV FRNT and an in-house DENV IgG assay, with a total of 36 monotypic DENV infections, 77 multitypic DENV infections, 56 primary ZIKV infections, 80 multi-flavivirus infections, and 151 unexposed (Table 2 and supplementary table 1). The four tests evaluated displayed a sensitivity near or above the ACIP recommendation (74.1%-91.2%); and high specificity (91.8%-98.6%); but only both versions of the CTK OnSite Dengue IgG rapid test reached the ACIP recommended target of ≥98% specificity (Table 3). The CTK OnSite rapid tests displayed a similar low proportion (∼1.8%) of false positives in the ZIKV primary and unexposed specimens. Of those two tests, only the CTK OnSite Dengue IgG rapid test R0065C with 76.2% sensitivity surpassed the ≥75% performance recommended by the ACIP. The CTK OnSite Dengue IgG Rapid Test R0065C-1.0 with equipment read yielded a sensitivity of 74.1% and nearly achieved the desired performance standards. The visual reads of the CTK OnSite Dengue IgG rapid test obtained by both laboratory technicians were identical for all specimens. Results showed that although the higher sensitivity (89.6% for R0065C, 81.9% for R0065C-1.0) of the rapid tests obtained when laboratory technicians performed the test read compared to the equipment reads, came at the cost of reduced specificity (95.7% for R0065C, 96.6% for R0065C-1.0) (Table 3). The ELISAs achieved 100% specificity in unexposed specimens but displayed some cross-reactivity against ZIKV. Although the Euroimmun anti-DENV NS1 Type 1-4 ELISA displayed a sensitivity (84.5%) high enough for pre-vaccination screening, it fell short on specificity (97.1%). A higher specificity was observed in the Euroimmun ELISA using NS1 than with the ELISA using VLPs as antigen that yielded a 91.8% specificity and sensitivity of 91.2%. (Table 3). Differences in sensitivity between tests were more pronounced in the detection of monotypic DENV specimens. Sensitivity for monotypic DENV was low to moderate (25.0-72.2%) for most tests. Multitypic DENV specimens and those with DENV and ZIKV exposure (multi-flavivirus) were detected with moderate to high sensitivity (≥83%) for all tests. The two CTK OnSite Dengue IgG rapid test versions showed a reduction in sensitivity when read by the manufacturer’s equipment compared to the visual read (Table 3). Because the Alta reader may not be available in all settings, we evaluated the performance of a combination of tests that could be used without the reader. We found that combining the Euroimmun anti-DENV NS1 Type 1-4 ELISA and the CTK OnSite R0065C read visually by the laboratory technician in a two-test algorithm, resulted in a specificity of 100% [98.2-100.0] and a sensitivity of 80.3% [74.0-85.7]; which meets the ACIP recommended performance criteria. Supplemental Figures 2-12 show flow diagrams for all test combinations indicated in Table 3.

**Table 3:**
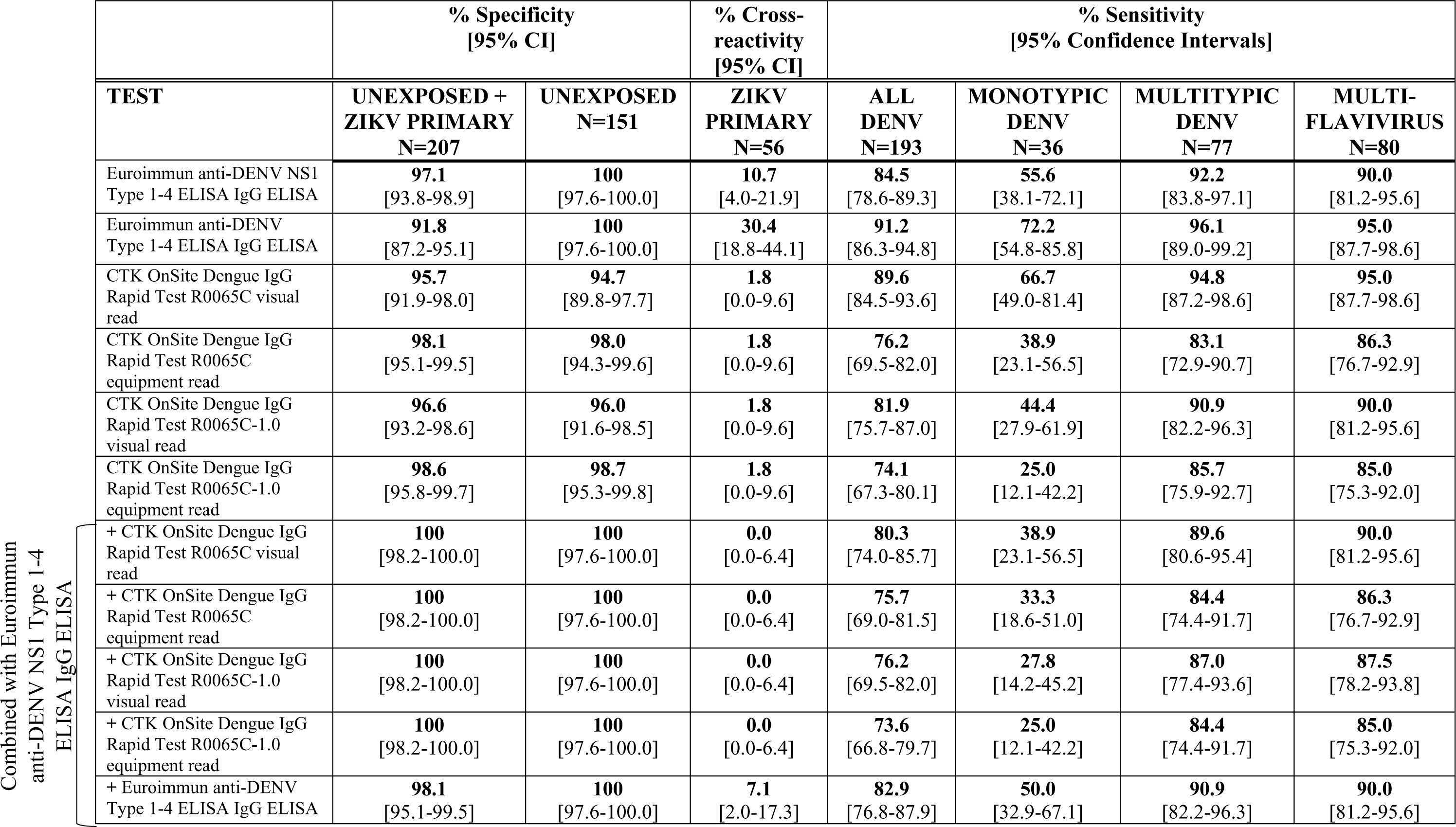
Specificity and sensitivity of commercial anti-DENV IgG tests used to determine previous dengue infection in 400 healthy individuals with specimens classified by virus neutralization and CDC DENV IgG ELISA.

### Additional evaluation of cross-reactivity

The current and future challenge of pre-vaccination screening arises largely because many dengue endemic countries also experienced ZIKV transmission. Therefore, to ensure dengue test specificity the Euroimmun anti-DENV NS1 Type 1-4 ELISA and CTK OnSite Dengue IgG rapid tests were evaluated with an additional 63 specimens from unexposed healthy individuals from Puerto Rico and well-characterized RT-PCR-confirmed past ZIKV infections from Nicaragua (Table 4).

**Table 4:**
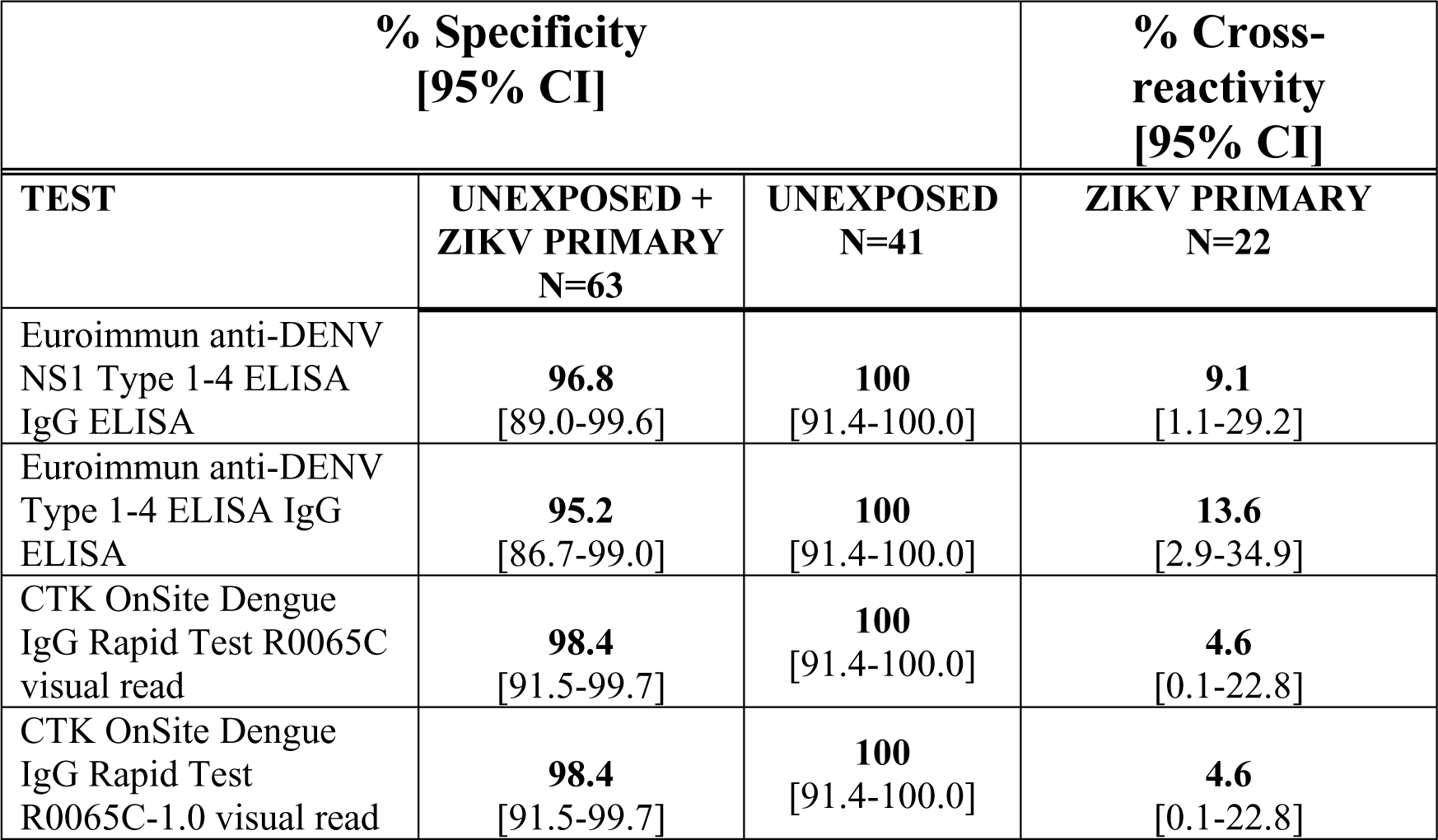
Specificity of high performing commercial anti-DENV IgG tests used to determine PDI with specimens from unexposed healthy individuals from Puerto Rico and past ZIKV infections from Nicaragua.

The highly specific Euroimmun anti-DENV NS1 Type 1-4 ELISA and CTK OnSite Dengue IgG rapid tests were evaluated with 12 samples from primary ZIKV specimens with high ZIKV neutralizing and binding antibody levels. The Euroimmun anti-DENV NS1 Type 1-4 ELISA IgG ELISA yielded 4/12 positives and the CTK OnSite Dengue IgG Rapid Tests both had 1/12 positives (Table 5). Both numbers are higher than expected and suggest that high ZIKV antibody levels could have an impact on test performance.

**Table 5.** Specimens with high ZIKV antibody tested in commercial anti-DENV IgG tests.

| Test | Cross-reactivity |
| --- | --- |
| Euroimmun anti-DENV NS1 Type 1-4 ELISA IgG ELISA | 4/12 (33%) |
| CTK OnSite Dengue IgG Rapid Test R0065C | 1/12 (8%) |
| CTK OnSite Dengue IgG Rapid Test R0065C-1.0 | 1/12 (8%) |

## Discussion

The FDA approval and subsequent ACIP recommendation of Sanofi Pasteur’s Dengvaxia^®^ has created a need for a highly accurate test for pre-vaccination screening. In the current study, we evaluated the performance of commercial anti-DENV IgG tests to determine which tests reached the ACIP’s required performance characteristics. Our study used well-characterized DENV and ZIKV specimens and included unexposed individuals from a dengue-endemic area, Puerto Rico, the United States territory with the largest number of dengue cases (4). The unbiased selection of specimens from healthy individuals should approximate the expected test performance in the community since most individuals experience asymptomatic DENV and ZIKV infections. As a reference standard, we used a combination of the CDC DENV IgG assay and DENV FRNT50. The correlation between the two assays was high with 399/400 specimens having concordant results. Our results showed that only the CTK OnSite Dengue IgG rapid test R0065C read with equipment reached the ACIP performance recommendation as a single test. Two tests were very close to meeting the recommendations but missed either for sensitivity (CTK OnSite Dengue IgG rapid test R0065C-1.0 with equipment read) or for specificity (Euroimmun anti-DENV NS1 Type 1-4 ELISA). Visual readings by the laboratory technicians resulted in increased CTK OnSite Dengue IgG rapid test sensitivity, but decreased specificity. Therefore, test interpretation must be determined by reading with the manufacturer’s equipment to ensure performance standards are maintained. In settings in the United States where the reader is not available, laboratory technicians could consider a visual read of the CTK OnSite Dengue IgG rapid test R0065C for pre-vaccination screening if a two-test algorithm is implemented with the combination of the Euroimmun anti-DENV NS1 Type 1-4 ELISA. Importantly, the effectiveness of screening is the same, regardless of whether the CTK OnSite Dengue IgG rapid test R0065C or the Euroimmun anti-DENV NS1 Type 1-4 ELISA is performed first. The combined use of these two tests resulted in the most sensitive and specific test algorithm that meets ACIP recommendations. Our data shows that there are commercial tests available for use as a pre-vaccination screening tool to confirm previous dengue infections and support safe vaccination practices (21). The CDC Dengue Branch has serum specimen sets for additional test evaluations and will provide updates of the results of these tests as they become available.

Our evaluation examined ZIKV cross-reactivity since it could interfere with pre-vaccination screening. False-positives in the CTK OnSite Dengue IgG rapid test R0065C occurred at the same frequency between ZIKV and non-ZIKV exposed individuals, but the Euroimmun anti-DENV NS1 Type 1-4 ELISA yielded more false-positives for those exposed to ZIKV than for those unexposed. We also performed limited testing with challenging ZIKV specimens that had high antibody binding and neutralizations titers that suggest there is a possibility of an increase in false positives in the event of ZIKV re-emergence.

Evaluations of commercial anti-DENV IgG tests have been limited by using convenient samples of well-characterized acute specimens from different cohorts that may not represent real-world scenarios (6). Moreover, a study evaluating commercial tests used for dengue diagnostics in Puerto Rico suggest they would not meet ACIP recommendations (8). Another study, which utilized data on virologically confirmed dengue infections collected before the Zika epidemic, found that the sensitivities of rapid diagnostic tests (RDTs) (40-70%) were lower than those of ELISAs (≥90%). However, consistent with our observations, ELISAs yielded a higher number of false positives than RDTs in specimens with ZIKV or flavivirus exposure. (7). Another factor that influences differences between the studies are the variance in the respective FRNT comparator such as assay conditions, virus strains, and cut-off (FRNT50 vs. FRNT90). Only few studies evaluated flavivirus cross-reactivity and did so separately from the specificity estimates. Studies on samples from non-ill individuals show high sensitivity (>90%), but inadequate specificity (<93.4%) for pre-vaccination screening. Most of the false positives were due to cross-reactivity with other flaviviruses (10, 11).

Our study is not without limitations. Due to the low number of monotypic DENV specimens and the low circulation of some DENV serotypes in Puerto Rico it was not possible for us to determine sensitivity by serotype. It is also possible that some of our specimens are misclassified. Our method of classifying specimens based on DENV IgG testing of acute (DPO 0-5) specimens was shown to correctly classify specimens approximately 85% of the time (22). We could not definitively classify DENV specimens as primary and secondary DENV in healthy individuals in the COPA cohort. Therefore, the impact of vaccination could be underestimated if only monotypic DENV specimens were classified as primary DENV cases. Some of the multitypic DENV specimens could indeed be primary DENV cases due to the cross-reactivity between the DENV serotypes. The cross-reactivity could also cause the misclassification of a small number of primary ZIKV specimens as multi-flavivirus and thereby result in a reduction of test sensitivity. Lastly, we limited the number of tests evaluated to those with publicly available information on test performance. It is possible that there are tests we did not evaluate that could be used for pre-vaccination screening of previous dengue infection either alone or in combination with another test.

In summary, the CTK OnSite Dengue IgG rapid test R0065C with equipment read was the only test able to satisfy the ACIP recommendations for test performance to determine previous DENV infection. However, the performance of this test in a two-test algorithm with the Euroimmun anti-DENV NS1 Type 1-4 ELISA results in a performance that surpasses the minimum test recommendations and reduces the concern of vaccinating individuals without previous dengue infection while providing the benefits of vaccination, including decreased illness and hospitalizations from dengue in the population at risk.

## Data Availability

All data produced in the present work are contained in the manuscript.

## Acknowledgements

We are grateful to Dr. Luisa I. Alvarado, the SEDSS and COPA staff and study participants for their contribution to this study.

## Disclaimer

The findings and conclusions in this report are those of the authors and do not necessarily represent the views of the Centers for Disease Control and Prevention.

---

**Figure S1.**
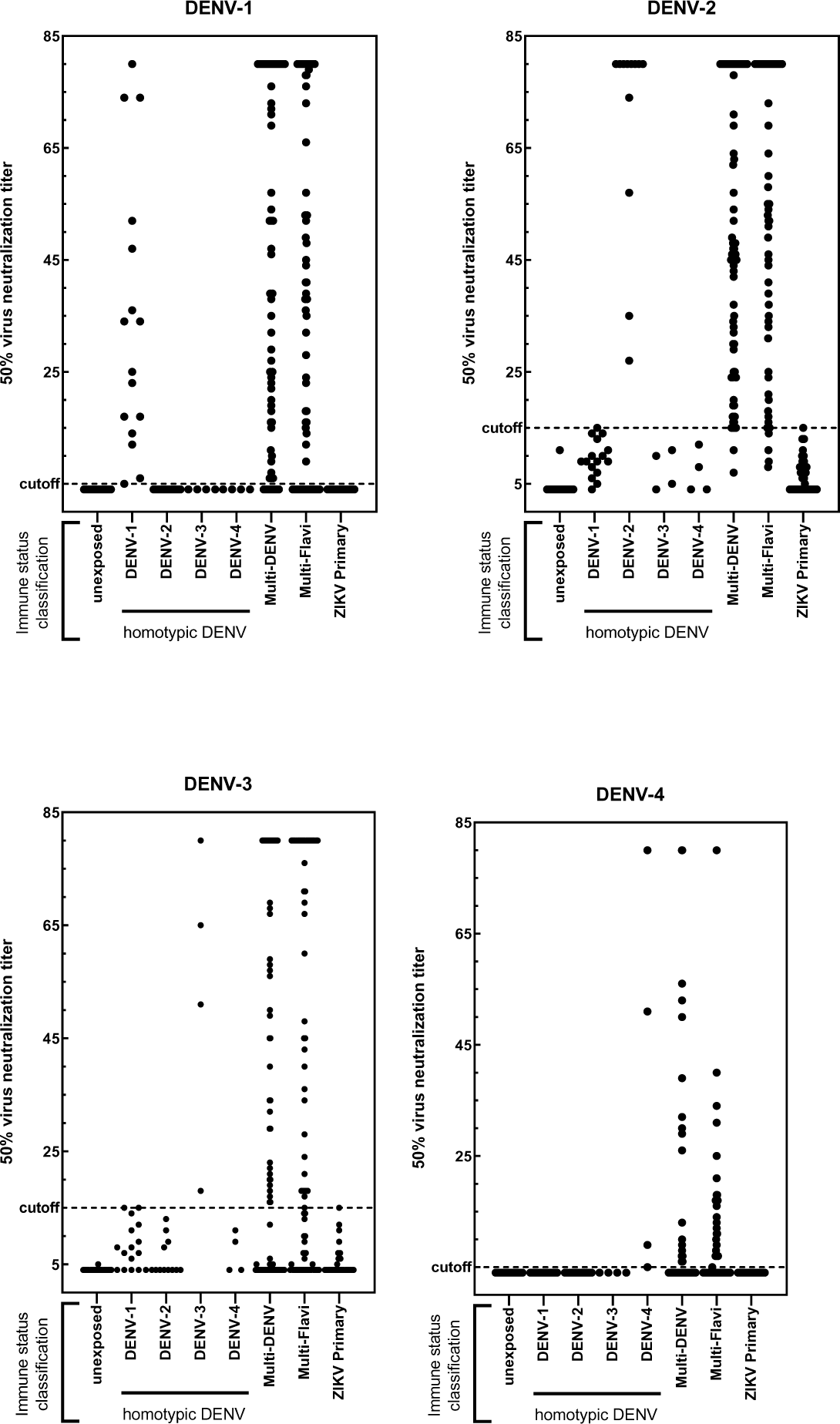
Sera from healthy children (ages 9-15) (n=400) living in Puerto Rico were tested in a focus reduction neutralization test and 50% virus neutralization titers (FRNT50) for DENV-1 (A), DENV-2 (B), DENV-3 (C), DENV-4 (D) and ZIKV (E) were determined as described in materials and shown according to immune status classification.

**Figure S2.**
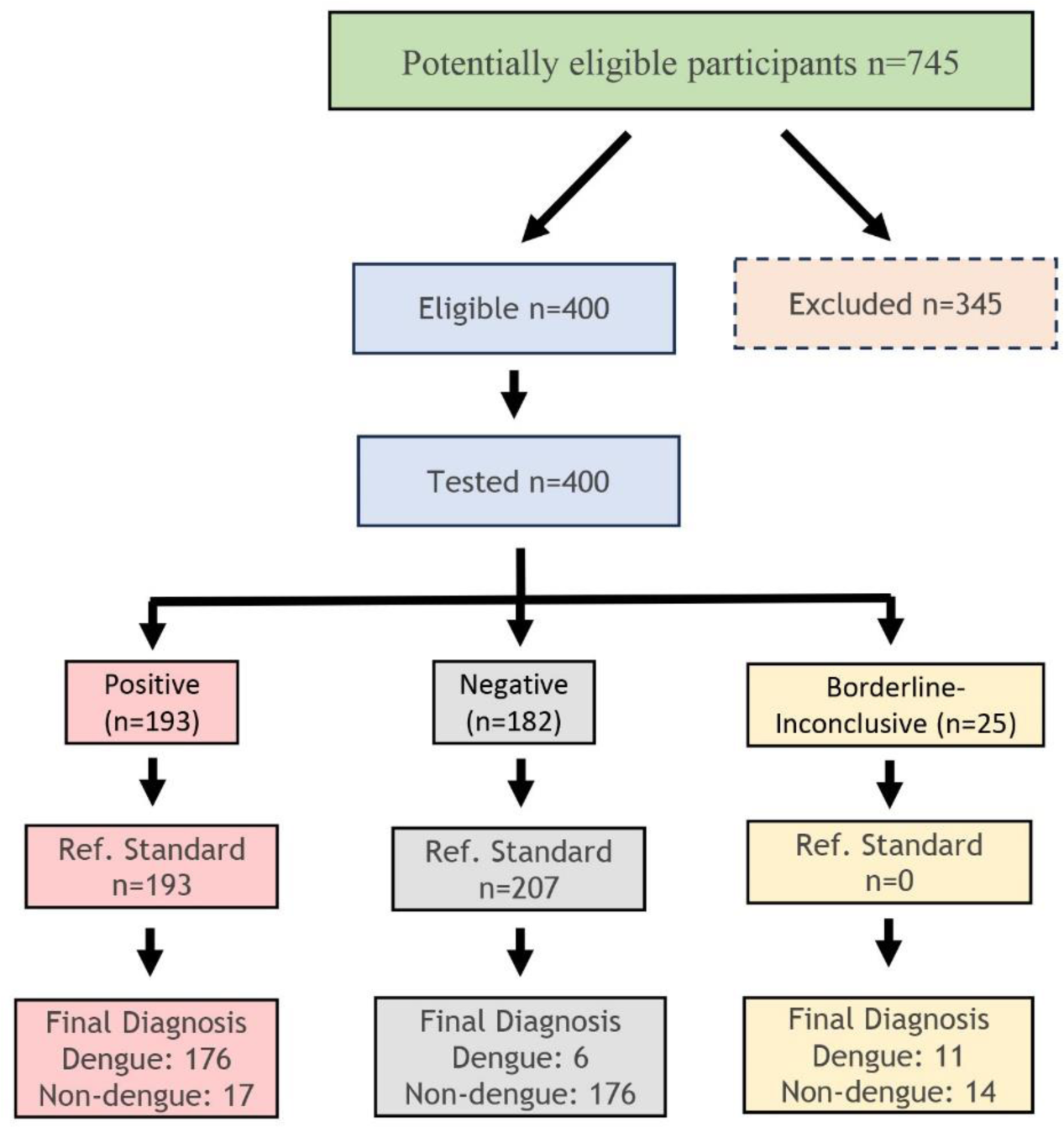
STARD flow diagram for Euroimmun anti-DENY Type 1-4 ELISA lgG ELISA

**Figure S3.**
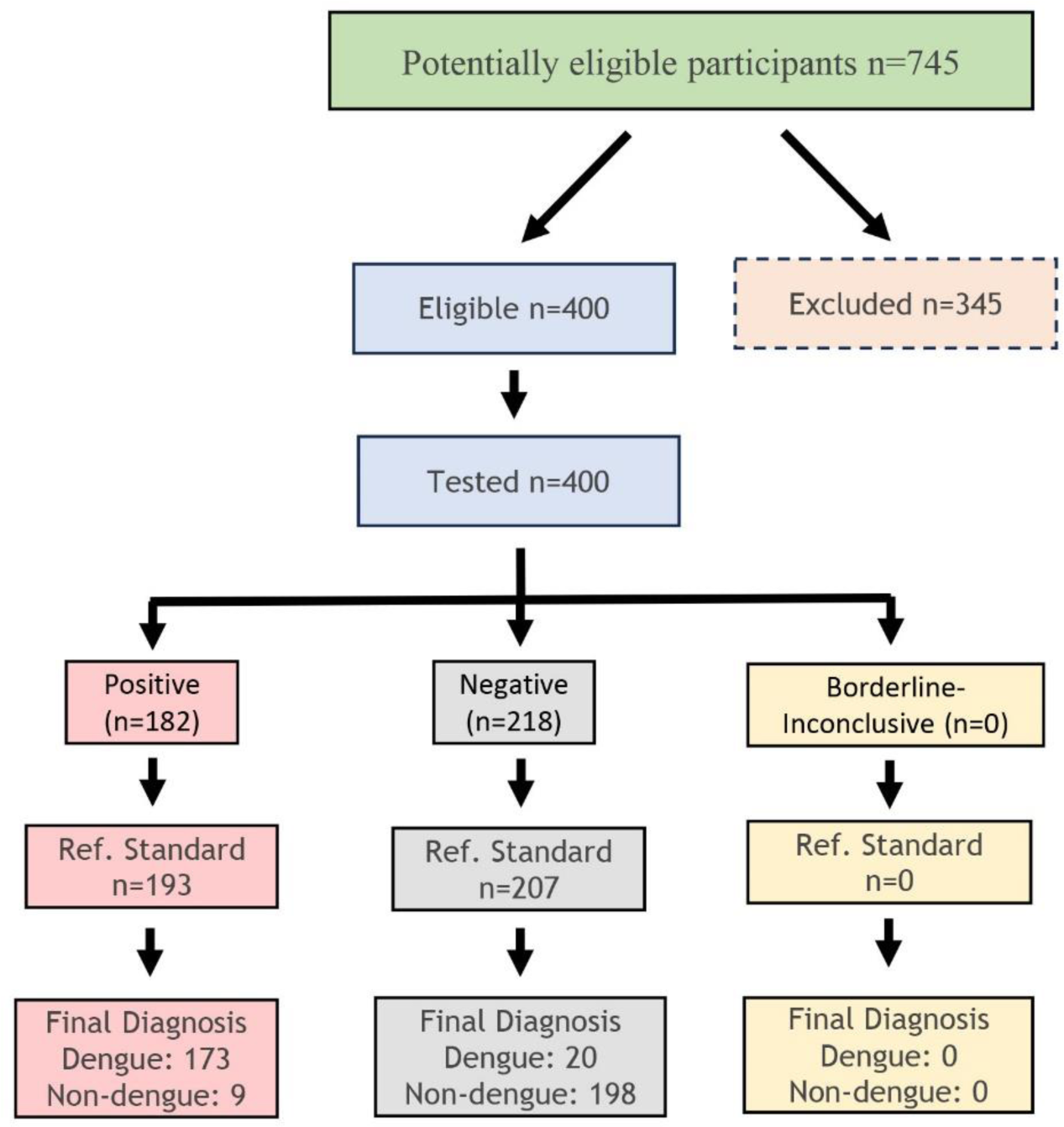
STARD flow diagram for CTK OnSite Dengue IgG Rapid Test R0065C visual read

**Figure S4.**
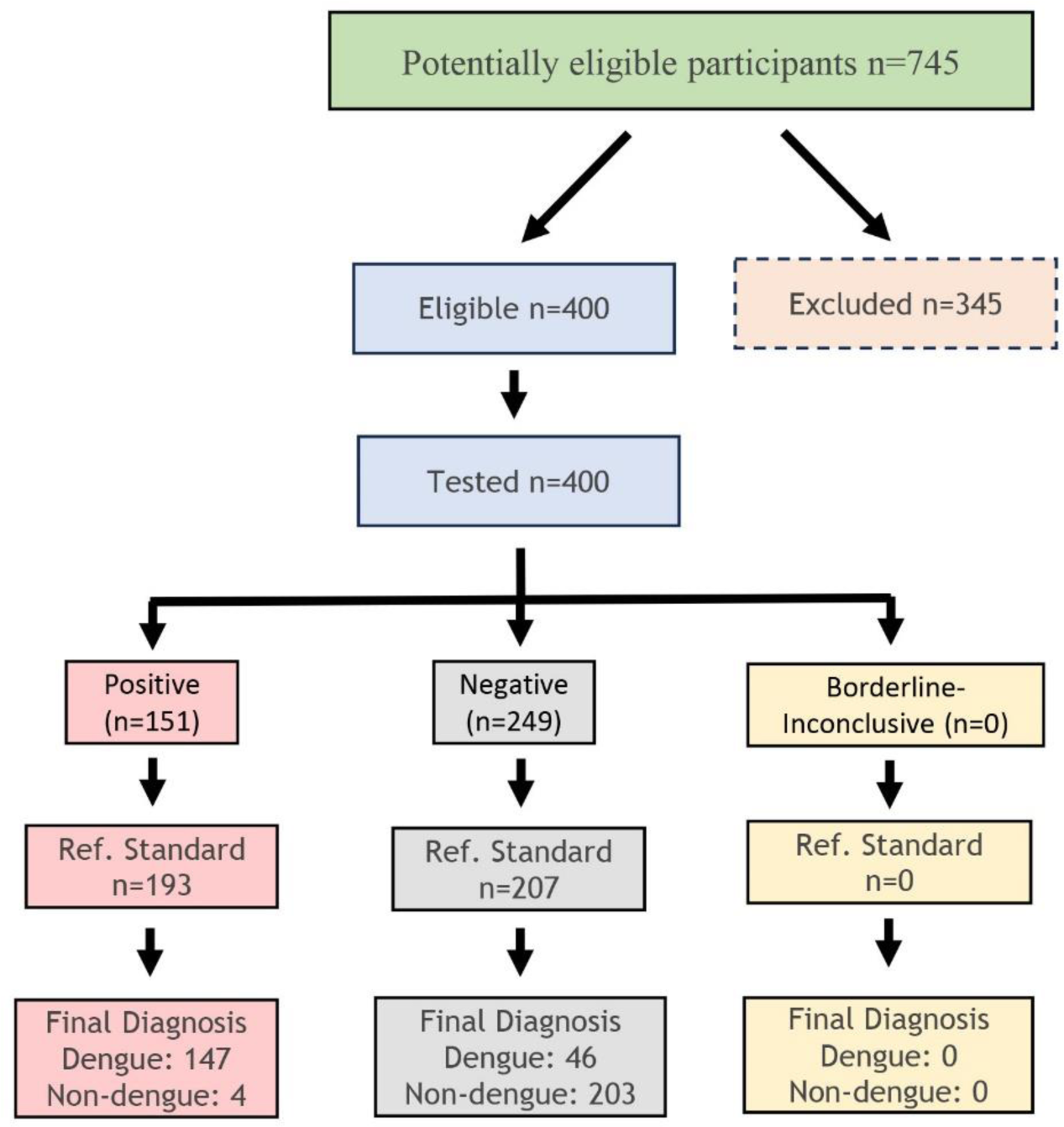
STARD flow diagram for CTK OnSite Dengue IgG Rapid Test R0065C equipment read

**Figure S5.**
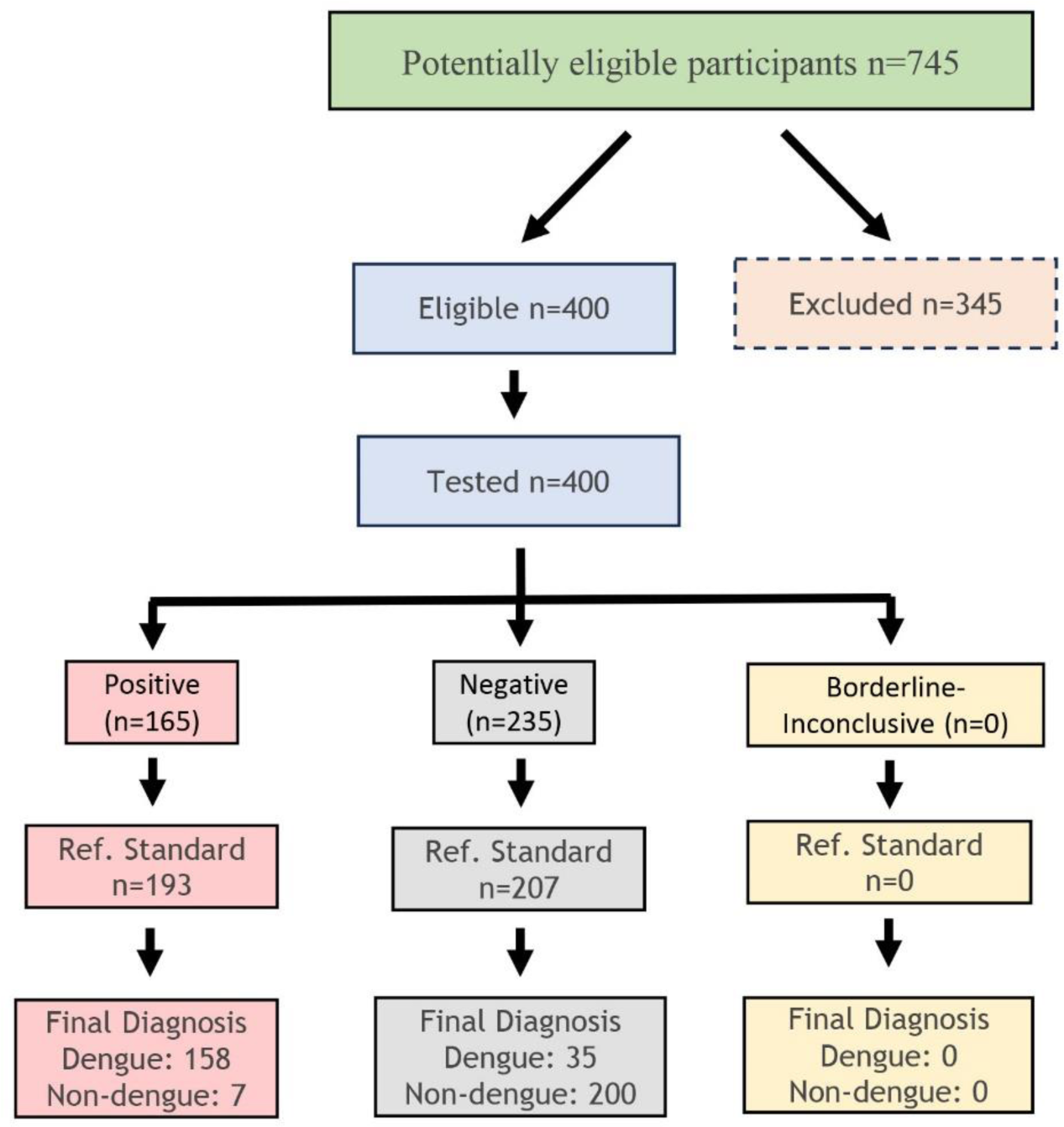
STARD flow diagram for CTK OnSite Dengue IgG Rapid Test R0065C-l.Ovisual read

**Figure S6.**
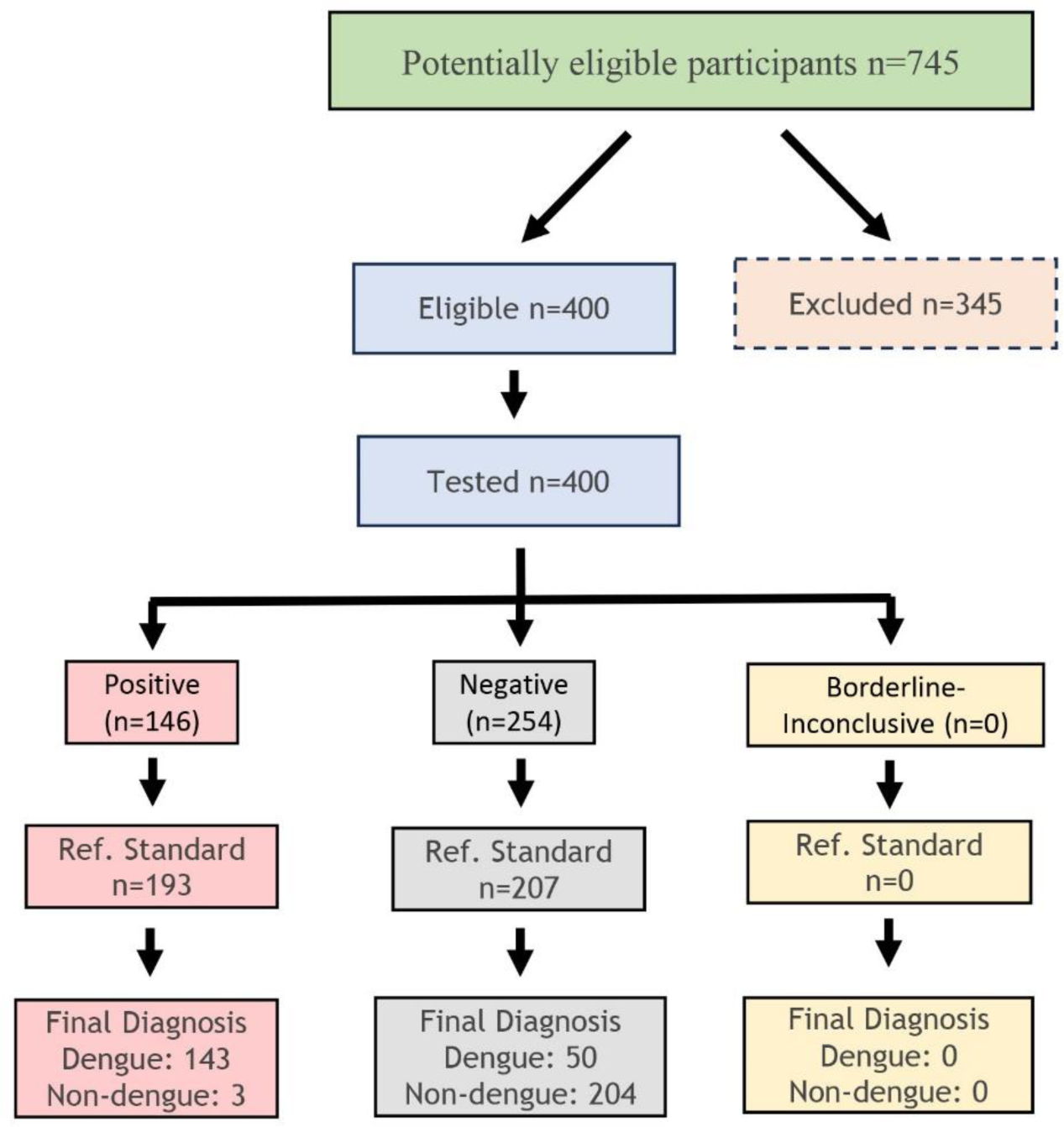
STARD flow diagram for CTK OnSite Dengue IgG Rapid Test R0065C-l.O equipment read

**Figure S7.**
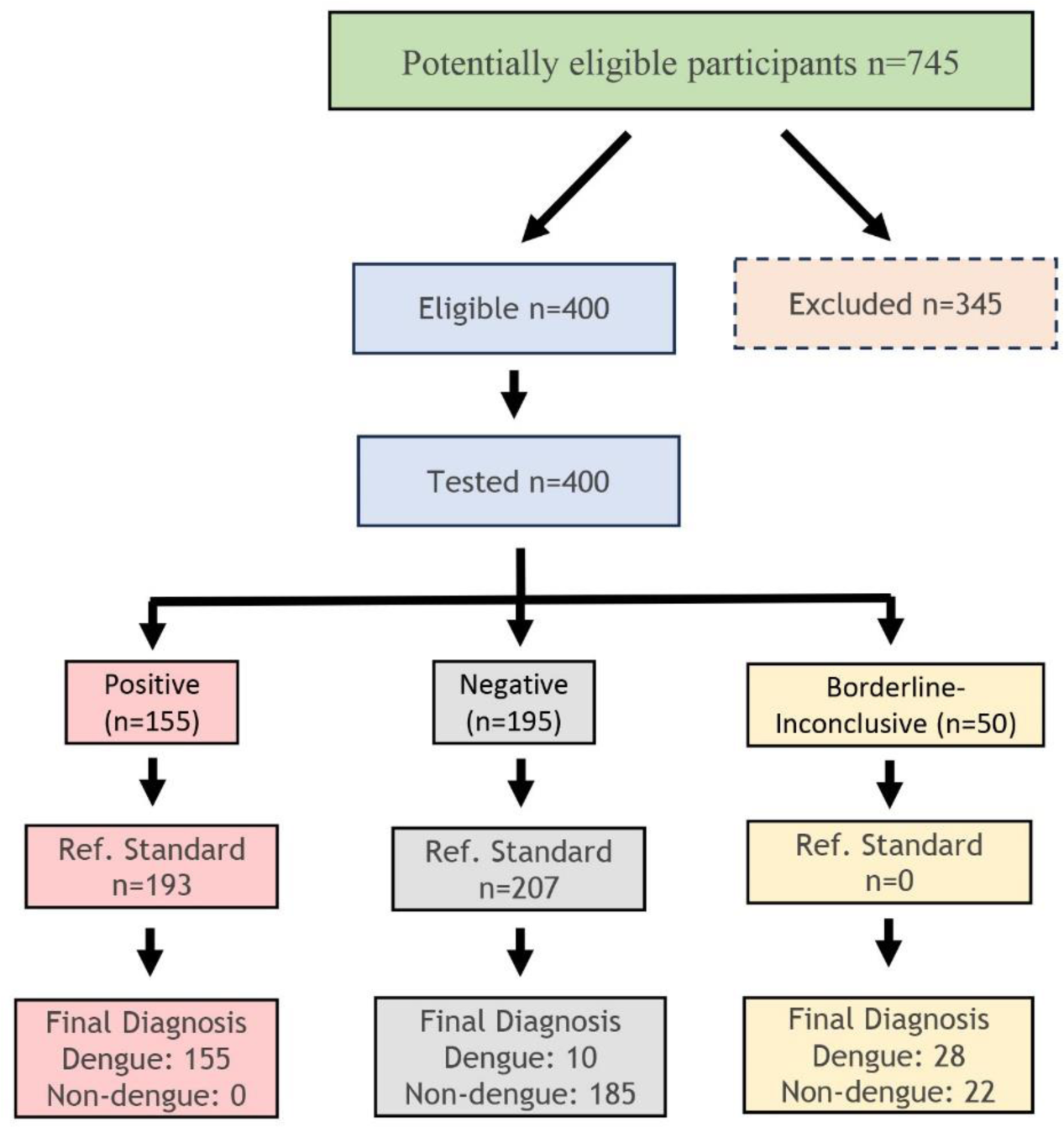
STARD flow diagram for Euroimmun DENY NSl Type 1-4 lgG ELISA+ CTK OnSite Dengue IgG Rapid Test R0065C visual read

**Figure S8.**
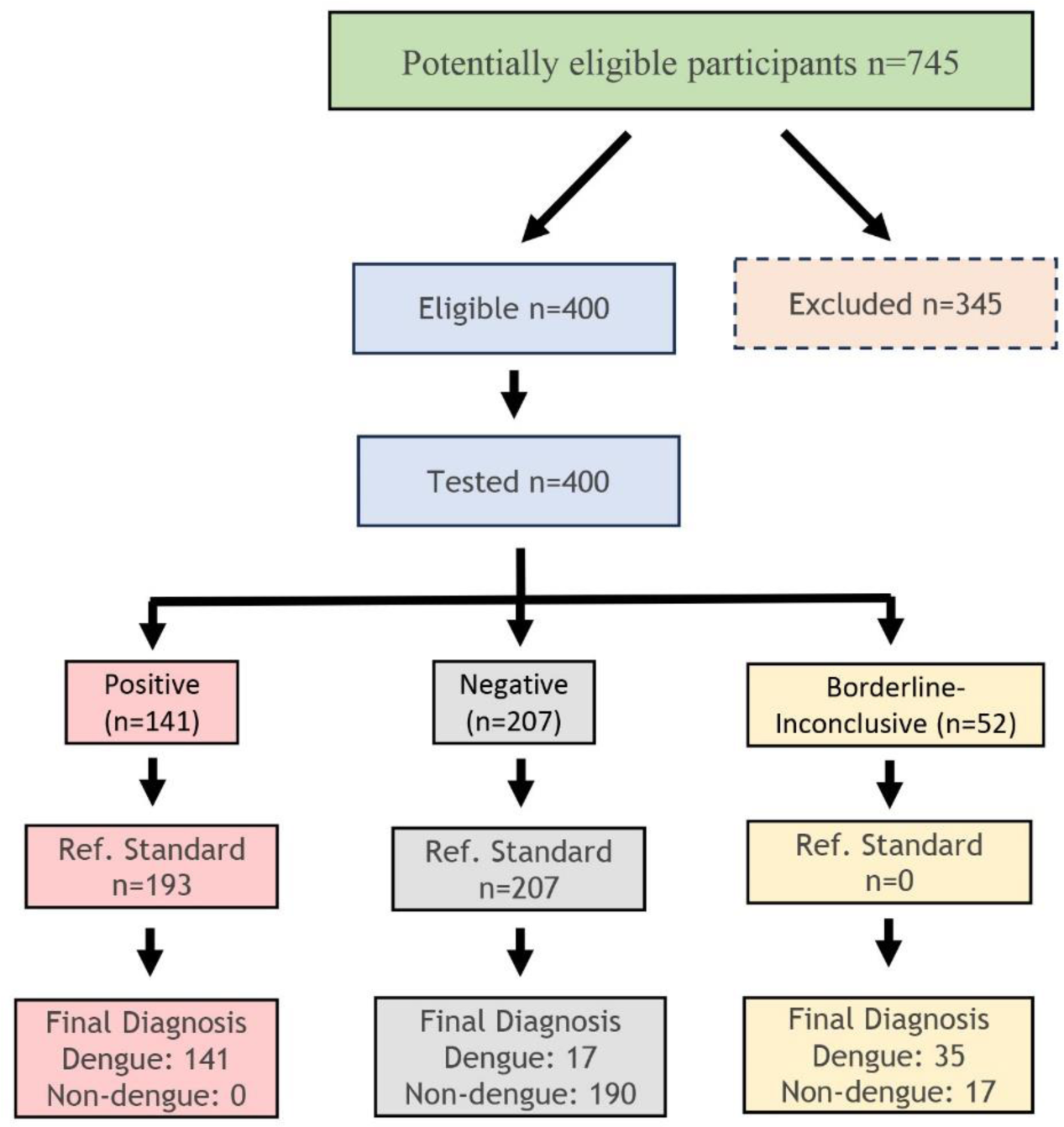
STARD flow diagram for Euroimmun DENY NSl Type 1-4 lgG ELISA+ CTK OnSite Dengue IgG Rapid Test R0065C equipment read

**Figure S9.**
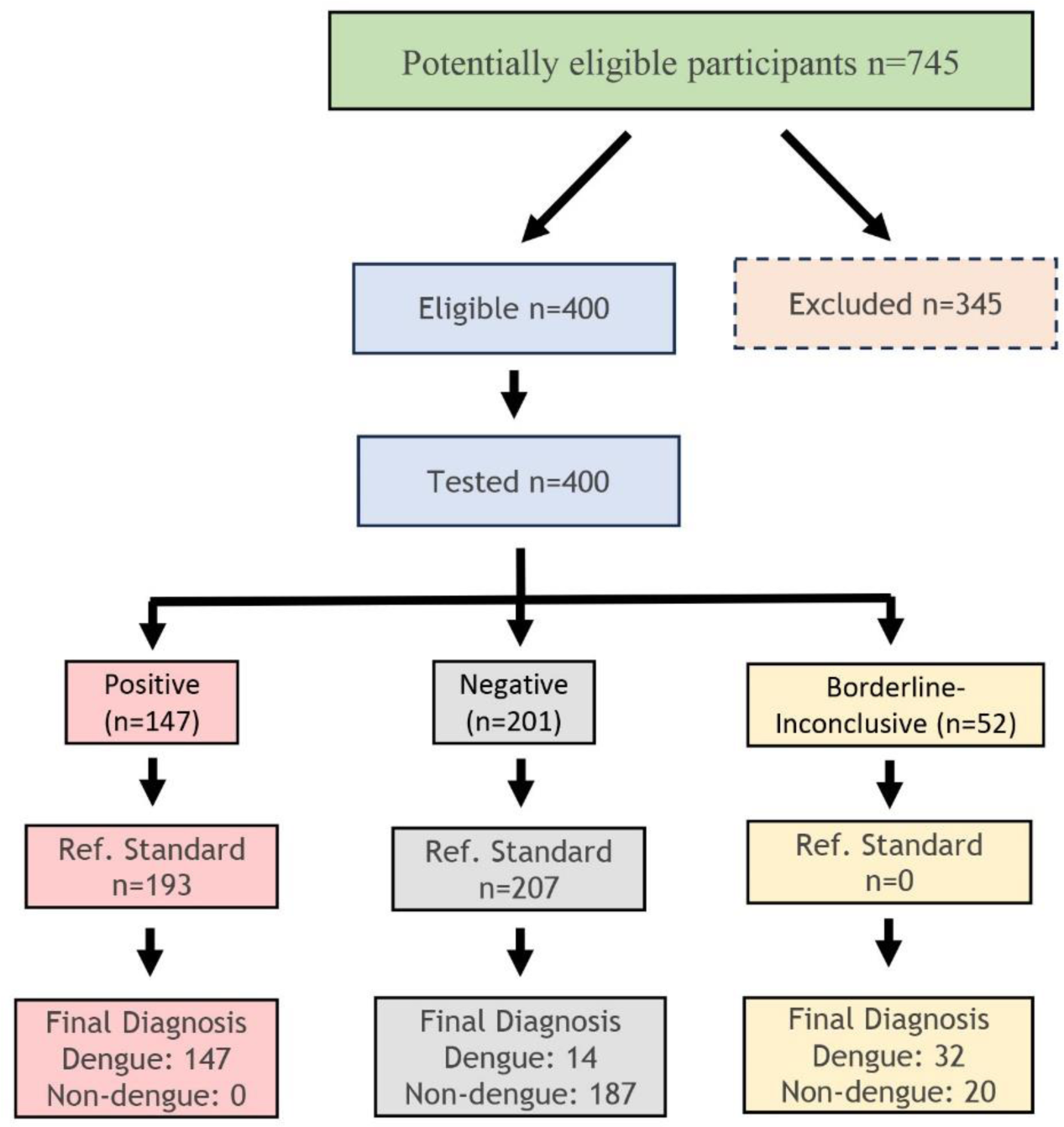
STARD flow diagram for Euroimmun DENY NSl Type 1-4 lgG ELISA+ CTK OnSite Dengue IgG Rapid Test R0065C-l.0 visual read

**Figure S10.**
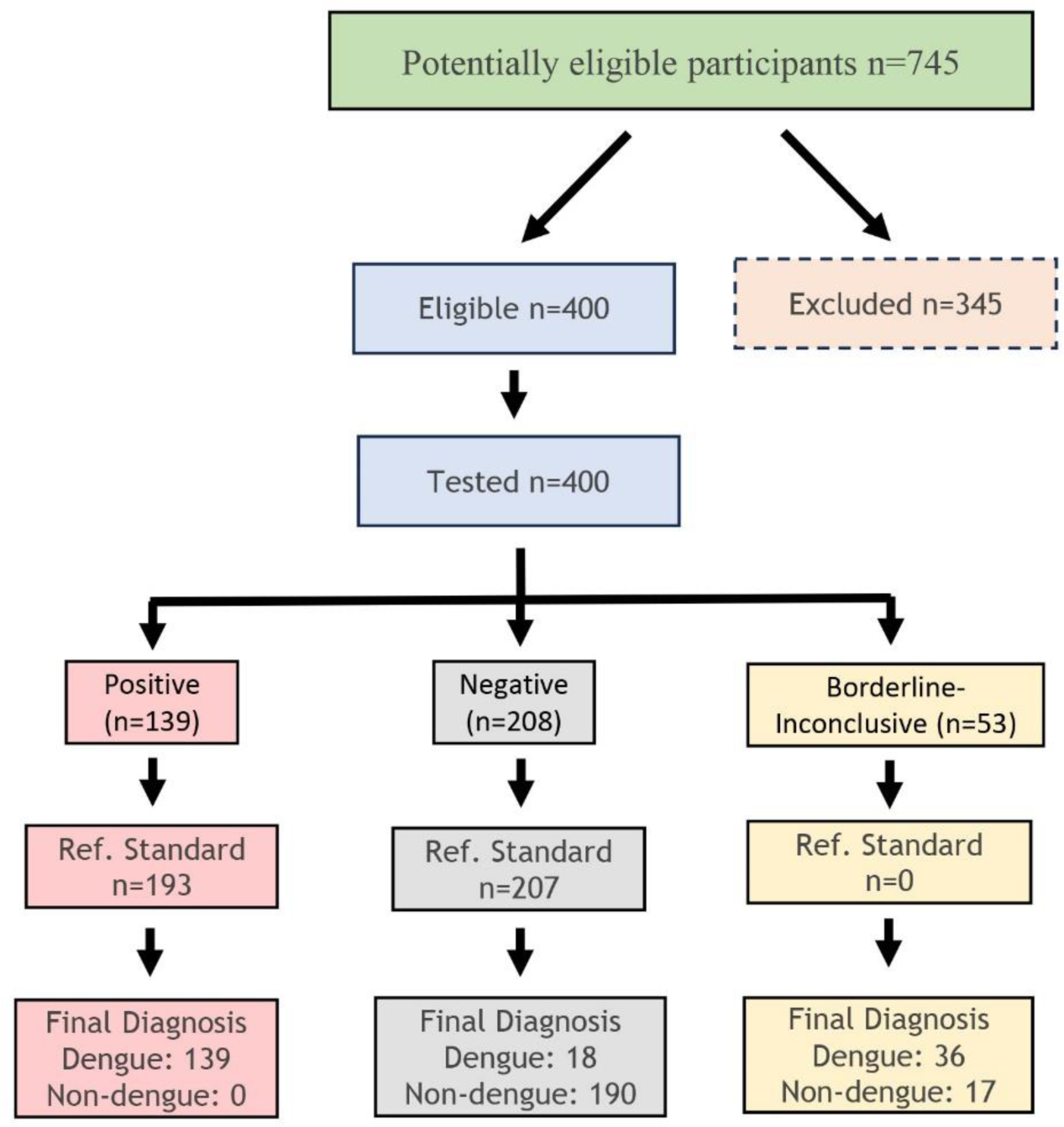
STARD flow diagram for Euroimmun DENY NSl Type 1-4 lgG ELISA+ CTK OnSite Dengue IgG Rapid Test R0065C-l.0 equipment read

**Figure S11.**
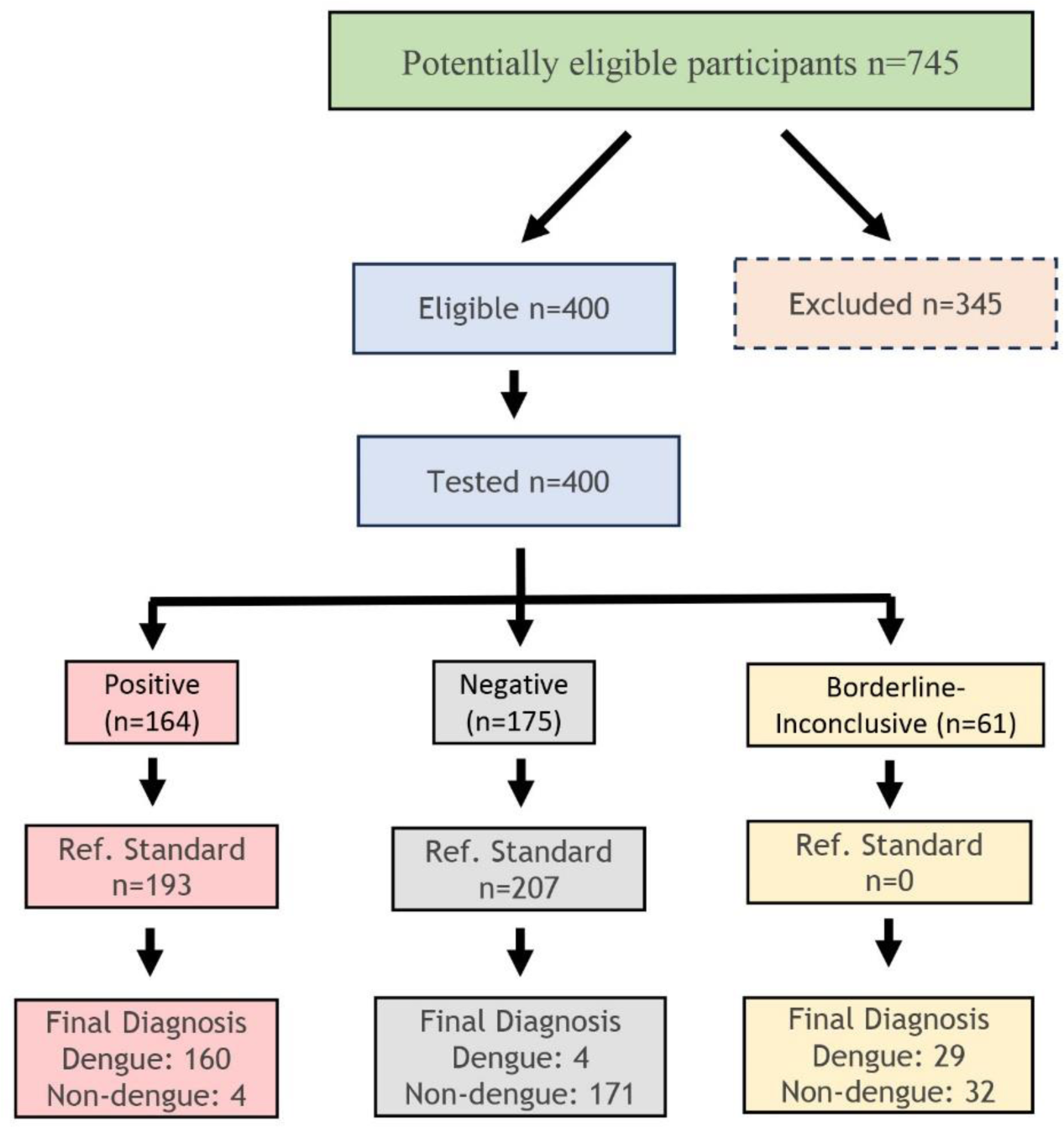
STARD flow diagram for Euroimmun DENY NSl Type 1-4 IgG ELISA+ CTK OnSite Dengue IgG Rapid Test R0065C visual read

**Figure S12.**
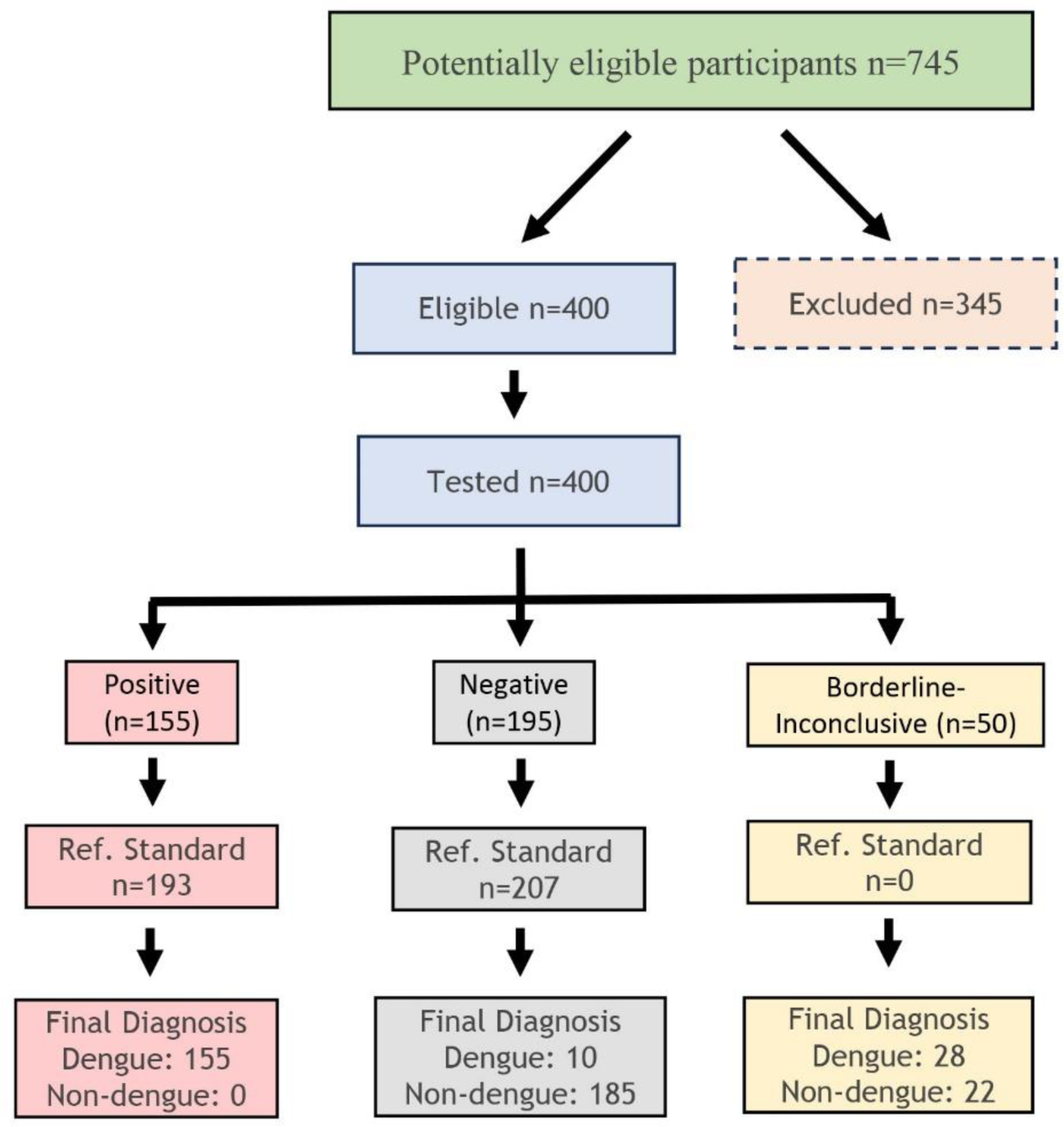
STARD flow diagram for Euroimmun DENY NSl Type 1-4 lgG ELISA+ CTK OnSite Dengue IgG Rapid Test R0065C visual read

## STARD checklist

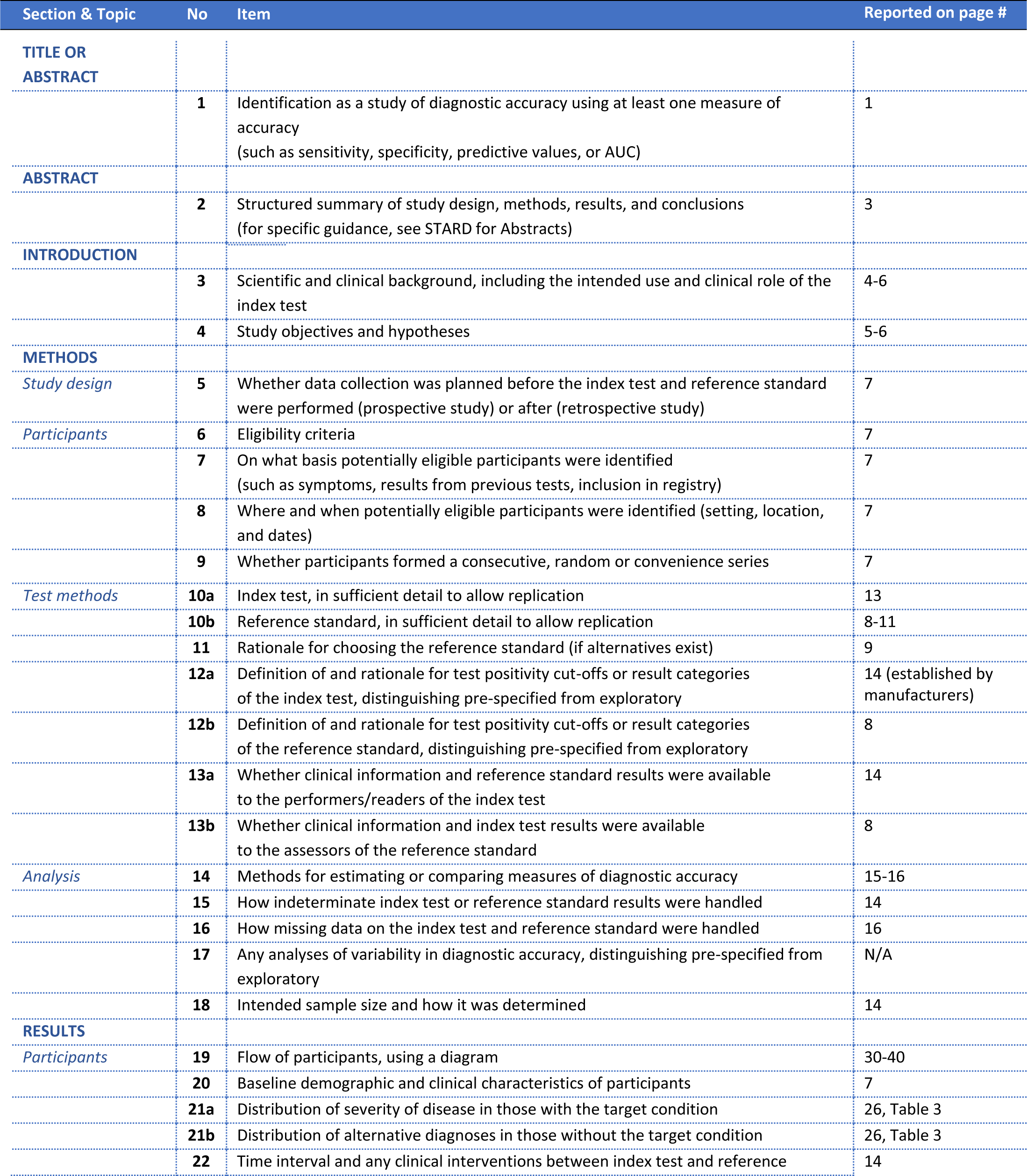
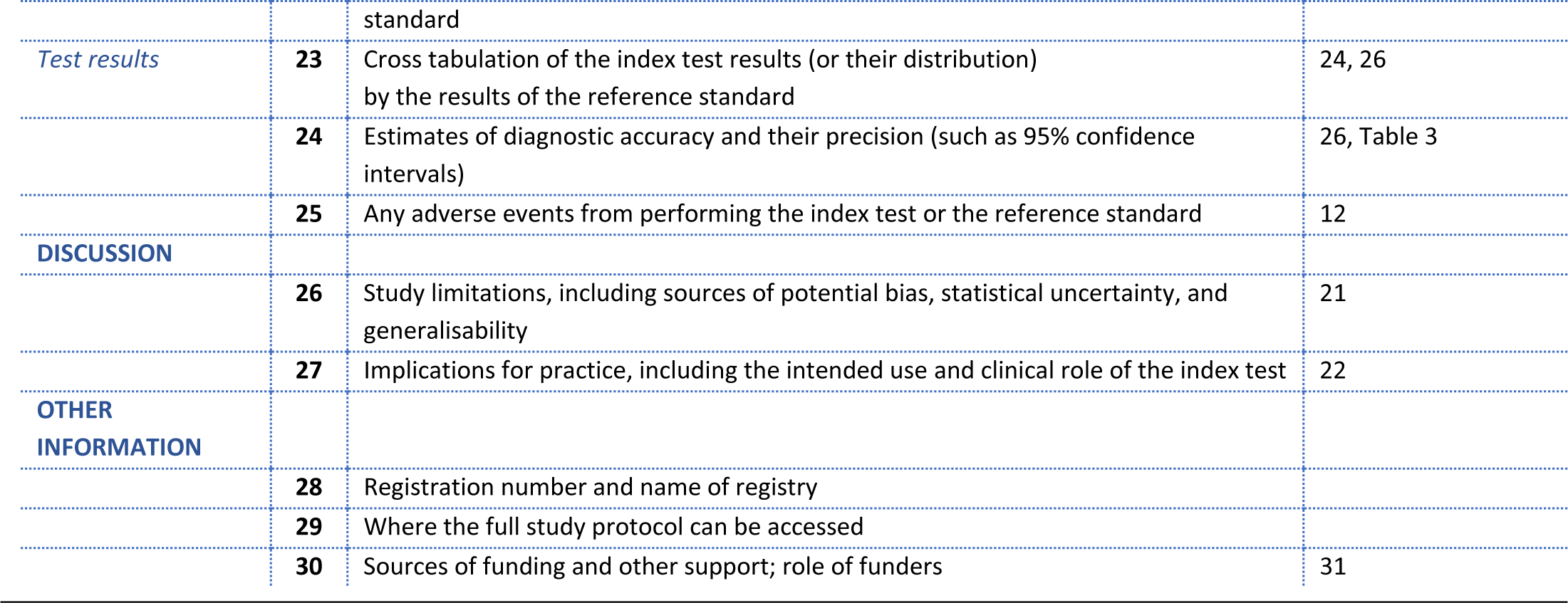

| Section & Topic | No | Item | Reported on page # |
| --- | --- | --- | --- |
| **TITLE OR ABSTRACT** |  |  |  |
|  | **1** | Identification as a study of diagnostic accuracy using at least one measure of accuracy (such as sensitivity, specificity, predictive values, or AUC) | **1** |
| **ABSTRACT** |  |  |  |
|  | **2** | Structured summary of study design, methods, results, and conclusions (for specific guidance, see STARD for Abstracts) | **3** |
| **INTRODUCTION** |  |  |  |
|  | **3** | Scientific and clinical background, including the intended use and clinical role of the index test | **4-6** |
|  | **4** | Study objectives and hypotheses | **5-6** |
| **METHODS** |  |  |  |
| *Study design* | **5** | Whether data collection was planned before the index test and reference standard were performed (prospective study) or after (retrospective study) | **7** |
| *Participants* | **6** | Eligibility criteria | **7** |
|  | **7** | On what basis potentially eligible participants were identified (such as symptoms, results from previous tests, inclusion in registry) | **7** |
|  | **8** | Where and when potentially eligible participants were identified (setting, location, and dates) | **7** |
|  | **9** | Whether participants formed a consecutive, random or convenience series | **7** |
| *Test methods* | **10a** | Index test, in sufficient detail to allow replication | **13** |
|  | **10b** | Reference standard, in sufficient detail to allow replication | **8-11** |
|  | **11** | Rationale for choosing the reference standard (if alternatives exist) | **9** |
|  | **12a** | Definition of and rationale for test positivity cut-offs or result categories of the index test, distinguishing pre-specified from exploratory | **14 (established by manufacturers)** |
|  | **12b** | Definition of and rationale for test positivity cut-offs or result categories of the reference standard, distinguishing pre-specified from exploratory | **8** |
|  | **13a** | Whether clinical information and reference standard results were available to the performers/readers of the index test | **14** |
|  | **13b** | Whether clinical information and index test results were available to the assessors of the reference standard | **8** |
| *Analysis* | **14** | Methods for estimating or comparing measures of diagnostic accuracy | **15-16** |
|  | **15** | How indeterminate index test or reference standard results were handled | **14** |
|  | **16** | How missing data on the index test and reference standard were handled | **16** |
|  | **17** | Any analyses of variability in diagnostic accuracy, distinguishing pre-specified from exploratory | **N/A** |
|  | **18** | Intended sample size and how it was determined | **14** |
| **RESULTS** |  |  |  |
| *Participants* | **19** | Flow of participants, using a diagram | **30-40** |
|  | **20** | Baseline demographic and clinical characteristics of participants | **7** |
|  | **21a** | Distribution of severity of disease in those with the target condition | **26, Table 3** |
|  | **21b** | Distribution of alternative diagnoses in those without the target condition | **26, Table 3** |
|  | **22** | Time interval and any clinical interventions between index test and reference standard | **14** |
| *Test results* | **23** | Cross tabulation of the index test results (or their distribution) by the results of the reference standard | 24, 26 |
|  | **24** | Estimates of diagnostic accuracy and their precision (such as 95% confidence intervals) | 26, Table 3 |
|  | **25** | Any adverse events from performing the index test or the reference standard | 12 |
| **DISCUSSION** |  |  |  |
|  | **26** | Study limitations, including sources of potential bias, statistical uncertainty, and generalisability | 21 |
|  | **27** | Implications for practice, including the intended use and clinical role of the index test | 22 |
| **OTHER INFORMATION** |  |  |  |
|  | **28** | Registration number and name of registry |  |
|  | **29** | Where the full study protocol can be accessed |  |
|  | **30** | Sources of funding and other support; role of funders | 31 |

